# Novel Coronavirus (COVID-19) Knowledge and Perceptions: A Survey of Healthcare Workers

**DOI:** 10.1101/2020.03.09.20033381

**Authors:** Akshaya Srikanth Bhagavathula, Wafa Ali Aldhaleei, Jamal Rahmani, Mohammadjavad Ashrafi Mahabadi, Deepak Kumar Bandari

## Abstract

**Background:** During the first week of March, the surge of coronavirus disease 2019 (COVID-19) cases reached over 100 countries with more than 100,000 cases. Healthcare authorities have already initiated awareness and preparedness activities beyond borders. A poor understanding of the disease among healthcare workers (HCWs) may result in delayed treatment and the rapid spread of infection. This study aimed to investigate the knowledge and perceptions of HCWs about COVID-19.

**Methods:** A cross-sectional, web-based study was conducted among HCWs about COVID-19 during the first week of March 2020. A 23-item survey instrument was developed and distributed randomly to HCWs using social media; it required 5 minutes to complete. A chi-square test was used to investigate the level of association among variables at the significance level of *p*<0.05.

**Results:** Of 529 participants, a total of 453 HCWs completed the survey (response rate: 85.6%); 51.6% were males, 32.1% were aged 25-34 years, and most were doctors (30.2%) and medical students (29.6%). Regarding COVID-19, most of the participants used social media to obtain information (61%), and a significant proportion of HCWs had poor knowledge of its transmission (61%) and symptom onset (63.6%) and showed positive perceptions of COVID-19 prevention and control. Factors such as age and profession were associated with inadequate knowledge and poor perception of COVID-19.

**Conclusion:** As the global threat of COVID-19 continues to emerge, it is critical to improve the knowledge and perceptions of HCWs. Educational interventions are urgently needed to reach HCWs beyond borders, and further studies are warranted.

## Introduction

Coronavirus (CoV) infections are emerging respiratory viruses and are known to cause illness ranging from the common cold to severe acute respiratory syndrome (SARS) [1]. CoV is a zoonotic pathogen that can be transmitted via animal-to-human and human-to-human interaction [2]. Multiple epidemic outbreaks occurred during 2002 (SARS), with ∼800 deaths, and 2012 (Middle East Respiratory Syndrome: MERS-CoV), with 860 deaths [2,3]. Approximately eight years after the MERS-CoV epidemic, the current outbreak of novel coronavirus (COVID-19) in Wuhan City, Hubei Province, China, has emerged as a global outbreak and significant public health issue [4]. On 30 January 2020, the World Health Organization (WHO) declared COVID-19 a public health emergency of international concern (PHEIC) [5]. Astonishingly, in the first week of March, a devastating number of new cases were reported globally, and COVID-19 emerged as a pandemic. As of 12 March 2020, more than 125,000 confirmed cases across 118 countries and more than 4600 deaths had been reported [6].

COVID-19 is spread by human-to-human transmission through droplet, feco-oral, and direct contact and has an incubation period of 2-14 days [6]. To date, no antiviral treatment or vaccine has been explicitly recommended for COVID-19. Therefore, applying preventive measures to control COVID-19 infection is the most critical intervention. Healthcare workers (HCWs) are the primary sector in contact with patients and are an important source of exposure to infected cases in healthcare settings; thus, HCWs are expected to be at high risk of infection. By the end of January, the WHO and Centers for Disease Control and Prevention (CDC) had published recommendations for the prevention and control of COVID-19 for HCWs [8,9]. The WHO also initiated several online training sessions and materials on COVID-19 in various languages to strengthen preventive strategies, including raising awareness and training HCWs in preparedness activities [10]. In several instances, misunderstandings among HCWs have delayed controlling efforts to provide necessary treatment [11], led to the rapid spread of infection in hospitals [12,13], and put patients’ lives at risk. In this regard, the COVID-19 epidemic offers a unique opportunity to investigate the level of knowledge and perceptions of HCWs during this global health crisis. In addition, we aim to explore the role of different information sources in shaping HCWs’ knowledge and perceptions of COVID-19 during this peak period.

## Methods

A prospective Web-based cross-sectional study was conducted using a survey instrument to obtain responses from HCWs globally during the first week of March 2020.

A 23-item survey instrument was developed using WHO course materials on emerging respiratory viruses, including COVID-19 [14]. The survey covered the domains of HCW characteristics, awareness, information sources, knowledge and perceptions related to COVID-19. The developed draft survey instrument was distributed to ten randomly selected faculty members to assess its readability and validity before pretesting among 20 randomly selected HCWs for clarity, relevance, and acceptability. Refinements were made as required to facilitate better comprehension and to organize the questions before the final survey was distributed to the study population.

### Content of the study tool

The survey instrument comprised 23 closed-ended questions and took approximately 3 minutes to complete. The 23-item questionnaire was divided into three parts, including participant characteristics (3 items), awareness of COVID-19 (2 items), source of information (4 statements/4-point Likert scale), knowledge about symptoms of COVID-19-affected patients (2 items), different modes of transmission (2 items), precautions and risk prevention (3 items) and perceptions of COVID-19 (7 items/true or false questions) [Supplementary file 1]. Sufficient time was given to participants to read, comprehend, and answer all the questions.

### Ethical considerations

Confidentiality of the study participants’ information was maintained throughout the study by making the participants’ information anonymous and asking the participants to provide honest answers. Eligible HCWs’ participation in this survey was voluntary and was not compensated. Informed consent was obtained from each participant prior to participation. The study was performed following the Helsinki Declaration as revised in 2013. The study was conducted following the Checklist for Reporting Results of Internet E-Surveys (CHERRIES) guidelines [15] [Supplementary file 2].

### Data analysis

The obtained data were coded, validated, and analyzed using SPSS version 24 (IBM, Armonk, NY, USA). Descriptive analysis was applied to calculate the frequencies and proportions. The chi-square test was used to investigate the level of association among variables. A *p*-value of less than 0.05 was considered statistically significant.

## Results

A total of 529 HCWs participated, 453 of whom completed the study questionnaire (85.6% response rate), including 234 (51.6%) men and 219 (48.3%) women; most of the participants were below 44 years of age (82.4%). The majority of participants were doctors (n=137, 30.2%) or medical students (n=134, 29.6%) and were from Asia (68%). Table 1 shows the sociodemographic characteristics of the participants. Almost all participants agreed that they had heard about COVID-19 (97.8%), but only 44.1% of them had the opportunity to attend lectures/discussions about COVID-19.

**Tables 1:**
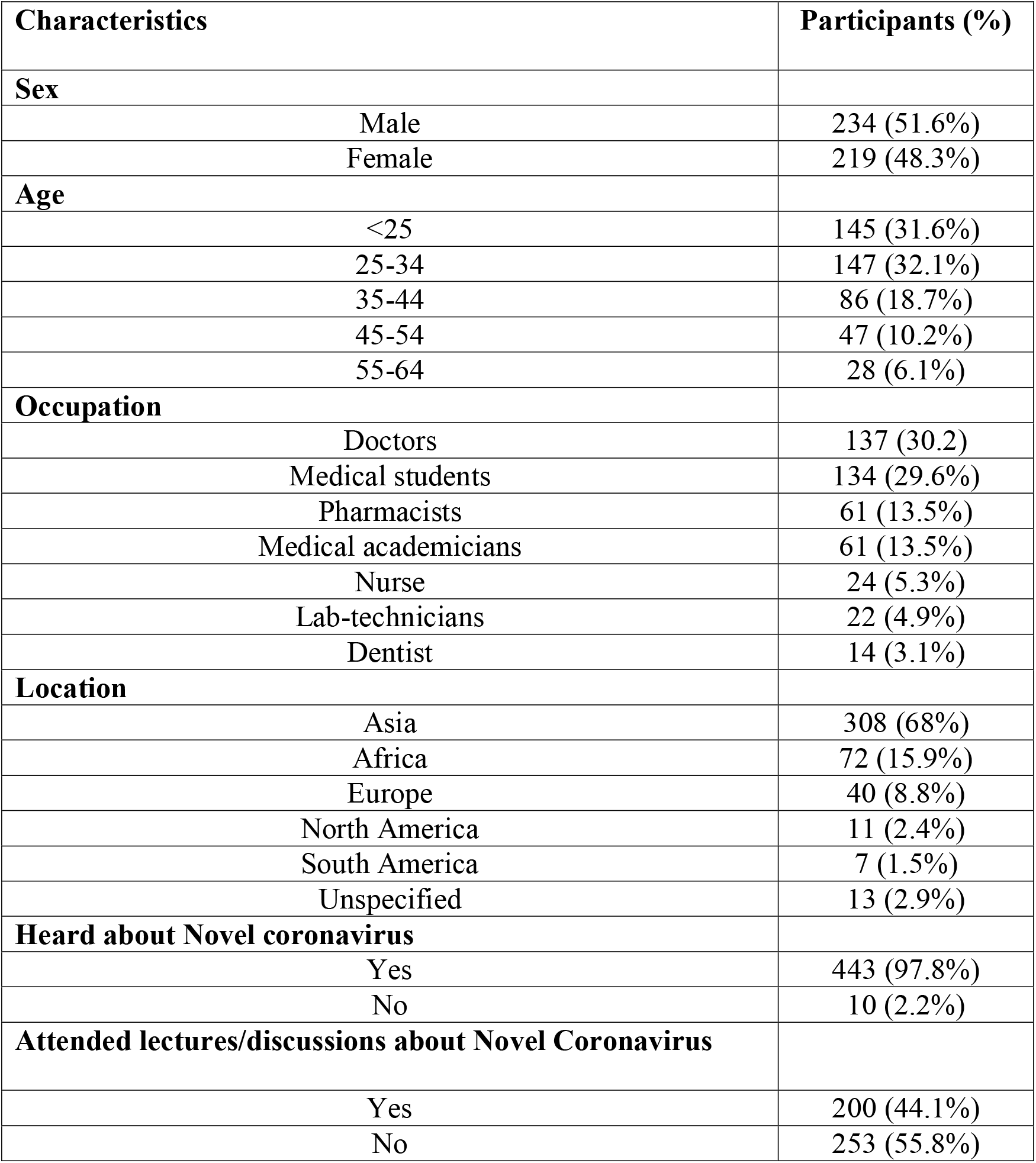
Sociodemographic characteristics of Healthcare workers’ (N=453)

| **Characteristics** | **Participants (%)** |
| --- | --- |
| **Sex** |  |
| Male | 234 (51.6%) |
| Female | 219 (48.3%) |
| **Age** |  |
| <25 | 145 (31.6%) |
| 25-34 | 147 (32.1%) |
| 35-44 | 86 (18.7%) |
| 45-54 | 47 (10.2%) |
| 55-64 | 28 (6.1%) |
| **Occupation** |  |
| Doctors | 137 (30.2%) |
| Medical students | 134 (29.6%) |
| Pharmacists | 61 (13.5%) |
| Medical academicians | 61 (13.5%) |
| Nurse | 24 (5.3%) |
| Lab-technicians | 22 (4.9%) |
| Dentist | 14 (3.1%) |
| **Location** |  |
| Asia | 308 (68%) |
| Africa | 72 (15.9%) |
| Europe | 40 (8.8%) |
| North America | 11 (2.4%) |
| South America | 7 (1.5%) |
| Unspecified | 13 (2.9%) |
| **Heard about Novel coronavirus** |  |
| Yes | 443 (97.8%) |
| No | 10 (2.2%) |
| **Attended lectures/discussions about Novel Coronavirus** |  |
| Yes | 200 (44.1%) |
| No | 253 (55.8%) |

### Source of information

When we asked about the participants’ source for reliable information about COVID-19, the primary sources of information about COVID-19 were official government websites and social media (Figure 1). Approximately 30% of the participants reported that they used news media (TV/video, magazines, newspapers, and radio) and social media (Facebook, Twitter, Whatsapp, YouTube, Instagram, Snapchat) to obtain information about COVID-19. Moreover, nearly 40% of the participants sometimes discussed COVID-19-related topics with family and friends.

**Figure 1:**
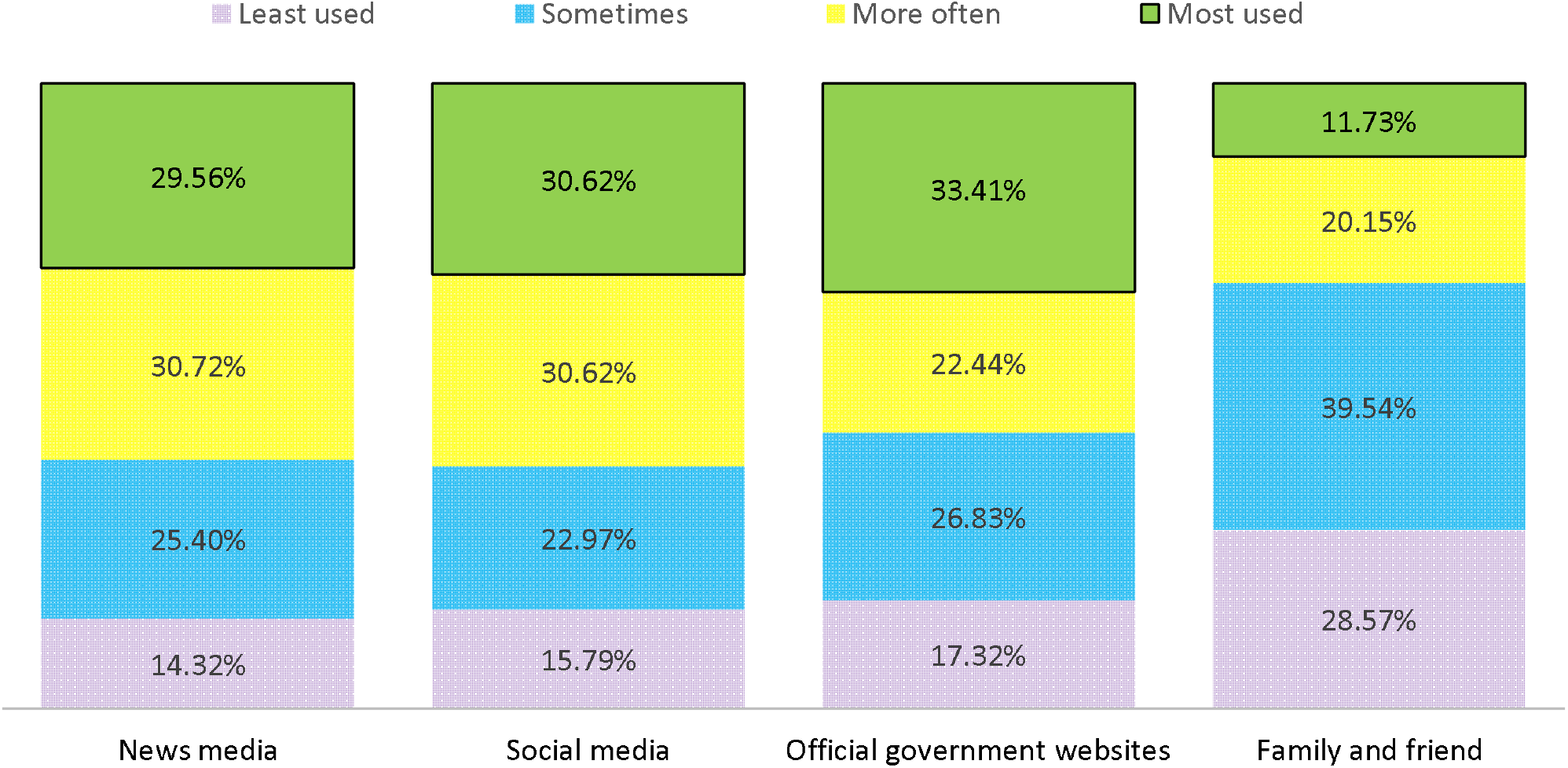
Source of Knowledge about Novel coronavirus

### Knowledge about COVID-19

Table 2 shows the knowledge about COVID-19 among HCWs. We identified significant knowledge gaps between doctors and other HCWs. For instance, approximately two-thirds of doctors and half of allied health workers thought that the origin of COVID-19 was bats (65.7% vs. 55.7%, *p*<0.05). A high majority of the HCWs (85.6%) agreed that maintaining hand hygiene, covering the nose and mouth while coughing, and avoiding sick patients could help to prevent COVID-19 transmission. A majority of the doctors agreed that COVID-19 could lead to pneumonia, respiratory failure, and death (84%, *p*<0.05) and that supportive care is the only treatment option that is currently available (83.2%, *p*<0.05). However, the participants’ knowledge about questions related to the mode of transmission and incubation period of COVID-19 was poor (*p*<0.05).

**Table 2:** Knowledge about Novel coronavirus (COVID-19) among Healthcare workers’ (N=453)

| Questions | Doctors<br>(n=137) | Allied health<br>workers (n=316) | Total Correct<br>responses | P-value |
| --- | --- | --- | --- | --- |
| COVID-19 is thought to be originated from bats | 90 (65.7%) | 176 (55.7%) | 266 (58.7%) | <0.05 |
| COVID-19 is transmitted through air, contact, fecal-oral routes | 50 (36.5%) | 127 (40.2%) | 177 (39%) | 0.46 |
| Headache, fever, cough, sore throat, and flu are symptoms of COVID-19 | 109<br>(79.6%) | 223 (70.6%) | 332 (73.2%) | <0.05 |
| The incubation period of COVID-19 (2-14 days) | 62 (45.3) | 103 (32.6%) | 165 (36.4%) | <0.05 |
| COVID-19 leads to pneumonia, respiratory failure, and death | 115 (84%) | 238 (75.3%) | 353 (77.9%) | <0.05 |
| Supportive care is the current treatment for COVID-19 | 114<br>(83.2%) | 193 (61%) | 307 (67.7%) | <0.05 |
| Hand hygiene, covering nose and mouth while coughing, and avoiding sick contacts can help in the prevention of COVID-19 transmission. | 117<br>(85.4%) | 271 (85.6%) | 388 (85.6%) | 0.96 |

### Perceptions about COVID-19

Over 78% of the HCWs exhibited a positive perception of COVID-19. A high majority of the HCWs knew that sick patients should share their recent travel history (92.7%), that flu vaccination is not sufficient to prevent COVID-19 (90.7%), and that COVID-19 is not fatal (88.5%). In addition, 87% felt that washing hands with soap and water could help to prevent COVID-19 transmission; 84.3% knew that symptoms appear in 2-14 days; and 85.6% agreed that all equipment used in wet markets should be cleaned every day. However, approximately 20% of HCWs were not clear about eating well-cooked meat during the outbreak [Table 3].

**Table 3:** Perception of Healthcare workers’ towards COVID-19

| Statements | Yes | No |
| --- | --- | --- |
| COVID-19 symptoms appear in 2-14 days | 382 (84.3%)* | 71 (15.6%) |
| COVID-19 is fatal | 52 (11.4%) | 401 (88.5%)* |
| Flu vaccinated is sufficient for preventing COVID-19 | 42 (9.2%) | 411 (90.7%)* |
| During the outbreak, eating well-cooked and safely handled meat is safe | 358 (78.1%)* | 95 (20.9%) |
| Sick patients should share their recent travel history with healthcare providers | 420 (92.7%)* | 33 (7.3%) |
| Disinfect equipment's and working area in wet markets at least once a day | 388 (85.6%)* | 65 (14.3%) |
| Washing hands with soap and water can help in prevention of COVID-19 transmission | 59 (13%) | 394 (87%)* |

\*correct answers

Items related to COVID-19-related perceptions among HCWs in the study were analyzed separately using the chi-square test to examine their association with age and sex and across different categories of people [Table 4].

**Table 4:** Association between respondents’ characteristics and perceptions of COVID-19

|  | Sex (%) |  |  | Age (%) |  |  |  | Profession (%) |  |  |  |
| --- | --- | --- | --- | --- | --- | --- | --- | --- | --- | --- | --- |
|  | Male<br>(234) | Female<br>(219) | P-value | <25<br>(145) | 25-44<br>(233) | 45-65<br>(75) | P-value | Doctors<br>(137) | Medical<br>students<br>(134) | Others<br>(182) | P-value |
| COVID-19 symptoms appear in 2-14 days |  |  | 0.85 |  |  |  | **<0.05** |  |  |  | **<0.05** |
| Yes† | 198<br>(84.6) | 183<br>(83.5) |  | 130<br>(89.6) | 207<br>(88.8) | 39<br>(52) |  | 126 (92) | 116<br>(86.5) | 146<br>(80.2) |  |
| No | 36 (15.3) | 36 (16.4) |  | 15<br>(10.4) | 26<br>(11.1) | 36<br>(48) |  | 11 (8) | 18 (13.5) | 36<br>(19.8) |  |
| COVID-19 is fatal |  |  | 0.19 |  |  |  | 0.78 |  |  |  | 0.31 |
| Yes | 22 (9.4) | 29 (13.2) |  | 127<br>(87.5) | 207<br>(88.8) | 68<br>(90.6) |  | 116 (84.6) | 112<br>(83.5) | 143<br>(78.5) |  |
| No† | 212<br>(90.6) | 190<br>(86.7) |  | 18<br>(12.5) | 26<br>(11.1) | 7<br>(9.4) |  | 21 (15.4) | 22 (16.5) | 39<br>(21.5) |  |
| Flu vaccinated is sufficient for preventing COVID-19 |  |  | 0.94 |  |  |  | 0.07 |  |  |  | **<0.05** |
| Yes | 24 (10.2) | 22 (10.1) |  | 21<br>(14.5) | 19<br>(8.1) | 5<br>(6.6) |  | 18 (13.1) | 16 (12) | 36<br>(19.8) |  |
| No† | 210<br>(89.7) | 197<br>(89.9) |  | 124<br>(85.5) | 214<br>(91.9) | 70<br>(93.4) |  | 119 (86.9) | 118 (88) | 146<br>(80.2) |  |
| During the outbreak, eating well-cooked and safely handled meat is safe |  |  | 0.67 |  |  |  | 0.13 |  |  |  | **<0.05** |
| Yes† | 192 (82) | 183<br>(83.5) |  | 113<br>(77.9) | 200<br>(85.8) | 63<br>(84) |  | 114 (83.2) | 98 (73.1) | 136<br>(74.7) |  |
| No | 42 (18) | 36 (16.4) |  | 32<br>(22) | 33<br>(14.2) | 12<br>(16) |  | 23 (16.8) | 36 (26.9) | 46<br>(25.3) |  |
| Sick patients should share their recent travel history with healthcare providers |  |  | 0.84 |  |  |  | 0.51 |  |  |  | **<0.05** |
| Yes† | 228<br>(97.4) | 214<br>(97.7) |  | 141<br>(97.2) | 229<br>(98.2) | 72<br>(96) |  | 131 (95.6) | 124<br>(92.5) | 158<br>(86.8) |  |
| No | 6 (2.6) | 5 (2.3) |  | 4<br>(2.8) | 4<br>(1.8) | 3 (4) |  | 6 (4.4) | 10 (7.5) | 24<br>(13.2) |  |
| Disinfect equipment's and working area in wet markets at least once a day |  |  | 0.26 |  |  |  | 0.54 |  |  |  | 0.41 |
| Yes† | 205<br>(87.6) | 199<br>(90.8) |  | 131<br>(90.3) | 206<br>(88.4) | 64<br>(85.3) |  | 116 (84.6) | 117<br>(87.3) | 149<br>(81.8) |  |
| No | 29 (12.4) | 20 (9.2) |  | 14<br>(9.7) | 27<br>(11.6) | 11<br>(14.7) |  | 21 (15.4) | 17 (12.7) | 33<br>(18.2) |  |
| Washing hands with soap and water can help in prevention of COVID-19 transmission |  |  | 0.58 |  |  |  | 0.24 |  |  |  | 0.88 |
| Yes† | 204<br>(87.2) | 187<br>(85.3) |  | 120<br>(82.7) | 207<br>(88.8) | 65<br>(86.6) |  | 118 (86.1) | 116<br>(86.5) | 160<br>(87.9) |  |
| No | 30 (12.8) | 32<br>(13.6%) |  | 25<br>(17.3) | 26<br>(11.1) | 10<br>(13.4) |  | 19 (13.9) | 18 (13.5) | 22<br>(12.1) |  |

†Correct answers; Significant at P<0.05 (bolded)

Nearly 90% of the youngest participants (<25 years) and 92% of the doctors believed that the symptoms of COVID-19 appeared as early as 2 to 14 days; the differences among the respondent groups were statistically significant (*p*<0.05). Moreover, a significant proportion of the doctors perceived eating well-cooked/handled meat to be safe (83.2%) (*p*<0.05). Medical students were found to have the perception that flu vaccination is not sufficient to prevent COVID-19 transmission (88%, *p*<0.05). A large number of allied health workers incorrectly believed that it is not safe to eat well-processed meat during the COVID-19 outbreak (25.3%, *p*<0.05), that COVID-19 is fatal (21.5%), that there is a delay in symptoms (19.8%), and that flu vaccination is sufficient (19.8%; *p*<0.05) compared with other participants in the respective groups.

## Discussion

Currently, COVID-19 is a global topic of discussion in the media and among the public, especially among HCWs and patients. With the currently mounting COVID-19 transmission raising tensions for everyone, including health officials and health systems, an important question arises regarding how we manage information to help frontline HCWs in times of public health crisis. For this reason, we investigated HCWs’ knowledge and perceptions of the prevention and control of COVID-19 during a global epidemic.

Knowledge and perceptions of COVID-19 varied across different categories of HCWs. Our study revealed that HCWs have insufficient knowledge about COVID-19 but showed positive perceptions of the prevention of COVID-19 transmission. We also found that more than 33% of the HCWs used official government websites as a primary source of information about COVID-19. This indicates that the COVID-19-related updates posted online by official government health authorities had positive implications for improving HCWs’ knowledge levels. Relying on authentic sources is a key factor in believing transparent information about the emerging COVID-19 infection and is essential for HCWs’ preparedness and response. However, a finding of considerable concern is that 60% of HCWs used social media as a source of information. Currently, the vast diversity of information available through the Internet, including unverified malicious information, can spread quickly and can misguide HCWs. In particular, health authorities and scientists have warned that widespread misinformation about COVID-19 is a serious concern causing xenophobia worldwide [4,16-19]. In this regard, HCWs should carefully evaluate COVID-19-related information and should use scientific and authentic content as information sources.

The findings of this study suggest significant knowledge gaps between the amount of information available about COVID-19 and the depth of knowledge among HCWs, particularly about the mode of transmission and incubation period of COVID-19. Additionally, many allied health workers had inaccurate knowledge that COVID-19 can be treated with antivirals and that there is a vaccine available. This is unfortunate because the surge of COVID-19 is globally devastating, and a large number of resources are provided by healthcare authorities to educate HCWs and improve their knowledge about COVID-19. Therefore, our findings were disappointing. Greater encouragement from health authorities is needed to assimilate COVID-19-related knowledge among HCWs, including doctors.

Generally, most participants had a positive perception of the prevention and control of COVID-19. However, discrepancies were identified in the perceptions of different categories of HCWs. For instance, only half (52%) of the HCWs aged 45-65 years believed that the symptoms of COVID-19 appeared as early as 2 to 14 days (*p*<0.05), and more than a quarter of the medical students thought that eating meat during the outbreak was unsafe. Approximately 20% of allied health workers believed that the flu vaccine is sufficient for COVID-19 prevention. Finally, a vast majority of HCWs strongly agreed that maintaining hygiene activities, reporting recent travel history when individuals are sick, and cleaning the equipment used in wet markets are strongly recommended.

## Limitations

We used WHO training material for the detection, prevention, response, and control of COVID-19 to develop a validated questionnaire. The developed questionnaire was pilot tested, and open-ended questions were limited to reduce information bias.

However, this study has some limitations that should be considered. This is a cross-sectional study conducted online among HCWs during alarming cases reported globally in the first week of March 2020. In addition, the data presented in this study are self-reported and partly dependent on the participants’ honesty and recall ability; thus, they may be subject to recall bias. Finally, due to the four-week closure of higher educational institutions in the UAE during the COVID-19 outbreak [16], the institutional review board was not approached. Despite these limitations, our findings provide valuable information about the knowledge and perceptions of HCWs during a peak period of COVID-19.

## Conclusion

We identified a significant gap in the source of information, poor knowledge levels, and discrepancies in perceptions of COVID-19 among our study participants. As the global threat of COVID-19 continues to emerge, greater efforts through educational campaigns that target HCWs and the wider population beyond borders are urgently needed.

## Data Availability

All the materials are attached as supplementary and information related to the study are in the manuscript.

## Acknowledgment

We thank all the study participants for their voluntary participation and for providing essential information.

## Authors’ contributions

ASB designed the study, developed the questionnaire, collected the data, analyzed the data, and prepared the manuscript. WAA designed the questionnaire, conducted the pilot test, and conducted the literature review. MMJ and JR distributed the questionnaire and filtered and analyzed the data. All authors read and approved the final manuscript.

## Available data and materials

All materials are attached as supplementary materials, and information related to the study is in the manuscript.

## Consent for publication

Not applicable.

